# Analysis of Effective Dose at Computed Tomography in a Modern 64 slice Multidetector CT System in an Irish Tertiary Care Centre with Local and International Reference Standards

**DOI:** 10.1101/2020.04.08.20057059

**Authors:** Tadhg-Iarla Curran, Michael Maher, Patrick McLaughlin, Fíona Coffey, Siobhan O’Neill

## Abstract

**Background:** Computed tomography (CT), by its nature, imparts a high dose of ionising radiation to patients being imaged. Diagnostic Reference Levels (DRLs) are used extensively in research but are also used in practice to establish if local imaging protocols have been optimised.

**Aim:** To audit effective dose (ED) at CT for four commonly performed examinations performed with a modern multidectector 64 slice CT (MDCT) system at large tertiary care centre by comparison with current international reference standards

**Methods:** 800 consecutive studies of CT brain, cervical spine, thorax and abdomen-pelvis were examined. All patients were imaged on 64-slice MDCT with study-specific protocols. Dose descriptors were extracted from image dose reports. ED was calculated using dose-length product conversion coefficients for all patients. Data was collated with Irish national and local DRLs, and published international point values.

**Results:** ED range at CT abdomen-pelvis of 2.03-26.04 mSv (mean 8.041 mSv) was 28.5% above the national DRL (9 mSv) and 26% above the local DRL (9.75 mSv). ED at CT thorax of 2.46-10.09 mSv (5.061 mSv) was 3.5% greater than local DRL (8.4mSv) and 42.5% greater than the national DRL (5.46 mSv). ED at CT brain of 2.25-8.08 mSv (3.075 mSv) was 100% above both the local (1.98 mSv) and national (2.06 mSv) DRLs. CT cervical spine ED of 1.49-21.94 mSv (5.14 mSv) was 79% greater than the local (3.07 mSv) and 100% greater than the national DRL (1.42 mSv).

**Conclusions:** All studies exceed national DRLs. A significant optimisation potential is noted, in particular, for studies involving the head and cervical spine. A further review of study protocols is necessary to facilitate optimisation of total study radiation dose.

## Section 1: Background and Literature Review

### Computed Tomography (CT)

The introduction of CT in the 1970s revolutionised radiological diagnosis and prompted a new era of medical imaging. Over time, there has been a significant increase in the use of the modality (1). In the US alone, the number of CT scans performed per annum has multiplied twentyfold, from 3.6 million scans performed in 1980 to 75 million scans performed today (1, 2). This figure continues to increase by 10% per annum as the applications for CT continue to diversify (2). The growth of CT practice worldwide has been driven by technological developments, most notably the introduction of helical CT scanning and multidectector CT systems which have improved the accessibility of CT to radiologists and patients, and the speed and quality of the resulting images (3).

The increased usage of CT as an imaging modality has not come without consequence. In comparison to conventional radiological techniques, CT imparts a relatively high radiation dose to the patient being imaged. CT alone accounts for over two-thirds of the total radiation dose from medical imaging in the US, but only 11% of all x-ray based procedures performed (1). In the UK, CT currently represents 9% of all radiological procedures and 47% of the medical radiation dose (4). This is in stark contrast to figures from 1989 which showed that CT represented only 2% of all radiological procedures but accounted for 20% of the cumulative radiation dose from diagnostic radiology (5).

Similar figures have been cited by the HSE in the first Irish survey of its kind on CT radiation dose in 2009. In Ireland, 211,000 scans are performed per annum (6). CT represents 10% of radiological examinations performed but 56% of the total radiation dose from medical imaging. Increases in the contribution of CT to radiation dose in Ireland have been noted in research conducted on Crohn’s disease from 1992 to September 2007 in one tertiary care centre. It was documented that CT accounted for 5.2% of imaging studies performed and 46.3% of radiation exposure between 1992 and 1997. However between 2002 and 2007 19.7% of imaging studies were CT scans, accounting for 84.7% of diagnostic radiation (7).

### Detrimental effects associated with CT

The detrimental effects associated with exposure to ionising radiation may be classified as either stochastic or deterministic. Deterministic effects occur above defined exposure thresholds, become worse with increasing exposure and include most types of direct tissue damage. Fortunately, they are rarely encountered with diagnostic imaging. Stochastic effects are those that occur without a dose threshold, but increase in probability with increasing dose (8). No safe level of exposure to radiation has been established and protracted exposure to low-level ionising radiation, including that for diagnostic use, could potentially be implicated in the induction of malignancies (2, 9). The overall risk of cancer from a single generic CT scan, extrapolated from studies conducted on atomic bomb survivors, is estimated to be in the range of 1.5 to 2% (10). Of the 70 million CT scans performed in the US in 2007, it is estimated that 29,000 cancer cases will result from CT-related ionising radiation exposure (11). Of 600,000 CT abdominal and CT head examinations performed per annum on children under the age of fifteen in the US, it is estimated that 500 fatal cancers are attributable to CT radiation (12).

### Relationship of excess radiation dose and image quality at CT

There is a unique relationship between radiation dose and image quality at CT. At low level radiation exposure there is a clear association between image quality and radiation dose however above a certain threshold this association is lost. This is due to an ‘uncoupling effect’ whereby the final image is divorced from radiation dose by digital and electronic manipulation (13). CT images always appear properly exposed at higher radiation doses as CT data is normalised to represent a fixed amount of attenuation relative to that of water (14). Therefore in contrast to conventional radiology, image quality is not impaired by overexposure and differences between an adequately exposed and overexposed image cannot be appreciated (2). As a result some organs may be overexposed with no increase in image quality while others may be underexposed with increased image noise and reduced image quality (2). Body habitus can have great effects on patient radiation dose due to the uncoupling effect. There has been a tendency to increase patient dose to avoid excessive noise in large patients and in thin section CT examinations. However technologists are not compelled to make corresponding reductions in the tube current–time product or the peak kilovoltage for scanning in small patients (14). This may lead to unnecessary overexposure and excessive radiation dose.

### Variability of radiation dose at CT

Radiation dose at CT is highly variable and a large disparity may exist in dose levels reported in different centres. This is to be expected due to different imaging protocols and the intrinsic differences between models and manufacturers of CT scanners (15). In a study of dose variations due to protocol differences alone, it was noted that even when using the same multidetector CT (MDCT) scanner model, a variation of up to sevenfold may exist in mean radiation dose for the same imaging procedure between centres (16). Intrinsic CT scanner-specific factors such as x-ray tube design, beam filtration, scanner geometry, anode angle and voltage waveform can lead to variations in CTDIvol of twofold for fixed tube current and kilovoltage settings (17). For all image types, MDCT scanners from the four most common manufacturers showed an average CTDIvol covariant variation across all organs of 34.1% (18). A fourfold range of dose values between the maximum and minimum radiation dose level in different hospitals may be noted for certain examinations (8).

Considerable variations in dose may exist also within a single radiology department due to scanner assignment. Most radiology departments have a variety of scanner models in use ranging from high-end MDCT scanners capable of state-of-the-art imaging modalities such as coronary angiography and virtual colonoscopy to older models designed more than a decade ago (19). Patients are exposed to up to three times less radiation when they undergo scans on 64 slice MDCT compared to older 4 and 16 slice MDCT scanners (19). There is relatively little research in the literature outlining the differences between single slice and multislice systems in terms of radiation dose. On average, MSCT tend to generate doses which are 34% greater than SSCT (20). The reasons for this have been attributed to more complex indications for MSCT scanning and injudicious use (21).

### Diagnostic Reference Levels (DRLs)

In order to optimise patient safety, X-ray examinations must fulfil two basic principles of radiation protection set by the International Commission on Radiological Protection (IRCP): examination justification and optimisation in accordance with the ALARA (as low as reasonably achievable) principle (22). It is now recognised that the implementation and use of DRLs is an efficient tool in the optimisation of diagnostic X-ray examinations, including CT (23). The concept of DRLs at CT was first introduced by the IRCP in 1990. In practice, they are derived by determining the 75th percentile of the distribution of a defined dosimetric quantity for routine conditions. Values are specific to a country or region assessing local radiological practices and should be updated periodically as technology advances. DRLs allow comparison of performance between various types of X-ray equipment (23). These reference levels help radiologists and radiographers identify and optimise imaging protocols which provide unusually high patient doses (22).They are not intended to suggest an ideal dose for a particular procedure and they do not represent an absolute upper limit for dose (24). If doses are found to be higher than the established reference doses without a justifiable reason, it is recommended that investigations identify the cause and appropriate remedial action is taken (25). It is now a legal requirement for all EU member states to initiate the establishment and use of DRLs under the Medical Exposures Directive. Many country- or region-wide surveys have been published in the last twenty years, all stating various values for DRLs, representative of the state of practice at the time and the population studied.

A review of the literature underlines the need to audit radiation dose at CT. This is made clear by the recent increases in ionising radiation exposure due to CT, the wide variations in radiation dose due to protocol and scanner-dependent factors, and the relative difficulty in managing radiation dose by image analysis alone. The application of national and local DRLs and the use of radiation doses published in international peer-reviewed journals may aid in both the standardisation and reduction of radiation dose at CT.

## Section 2: Aims and Objectives

### Aim

- To audit effective dose at CT for four commonly performed examinations performed with a modern multidectector 64 slice CT system at large tertiary care centre by comparison with current international reference standards.

### Objectives

This aim will be met by carrying out the following objectives:

- Applying for ethical approval from the Clinical Research Ethics Committee of the Cork Teaching Hospitals.
- Identifying a suitable sample of CT examinations from a single CT scanner with which to measure effective dose.
- Constructing a bespoke database of data obtained.
- Selecting data from the literature for formation of a reference interval database and creating a chronological framework of this data to ensure accurate comparison with current results.
- Examining dose reports from all members of the population for key parameters including patient medical registration number, patient date of birth, scan range, CTDIvol and DLP.
- Examining the literature and selecting up to date *E*_*DLP*_ conversion coefficients which reflect the recent changes in weighting factors in the IRCP 103 guidelines.
- Calculating the effective dose for all scan types involved using Microsoft Excel 2007 calculation software by application of the *E*_*DLP*_ coefficient to the DLP data obtained.
- Reviewing results with a qualified Specialist Registrar in Radiology for data cleaning.
- Performing statistical analysis of all data obtained in order to ensure accurate and succinct observations.

## Section 3: Methods

### 3.1 Obtaining the study sample

All study samples were obtained from a single CT scanner at CUH between 1 January 2010 and 23 October 2011 for this cross sectional study. 200 sequential studies were individually examined for four different scan types. This represented a total study sample of 800 scans. The four categories selected were CT abdomen and pelvis routine, CT thorax with contrast, CT cervical spine and non-contrast CT brain routine. These studies and the accompanying radiology dose reports were accessed using the CUH picture archiving and communication system (PACS), which uses IMPAX 6 software (Agfa Healthcare NV, Belgium). All data was exported to a HP Compaq Xeon dc5050 workstation (Hewlett-Packard Company, CA, US). A database was constructed using Microsoft Office Excel 2007 (Microsoft Corporation, CA, US) for collation of data.

### 3.2 Patient selection criteria

Patient lists were populated using advanced search methods on the PACS database. The search parameters used included the imaging station name, patient age at acquisition, study description and study date. The application of advanced search methods allowed more stringent control of the inclusion and exclusion criteria for the patient lists produced.

#### Inclusion criteria

- All scans were obtained from the General Electric Lightspeed VCT-XTE 64 slice multi-detector CT (MDCT) system. Scans obtained from the alternative Toshiba Aquilon II 4 slice MDCT (Toshiba Medical Systems Corporation, Tochigi, Japan) were representative of a less modern 4 slice MDCT system. Therefore these were not included.
- Patients were at least 18 years of age or older at the time of acquisition of the studies

#### Exclusion criteria

- Patients who underwent more than one helical scan (n=27)
- Patients under the age of 18 at the time of acquisition of the study (n=54)
- Patients who underwent CT brain with contrast and CT brain pre and post contrast (n=28)
- Scans not for clinical use (n=1)

### 3.3 Computed Tomography

All studies were performed on the General Electric LightSpeed VCT-XTE multi-detector 64 slice CT system (GE Healthcare, Wisconsin, US) with a 0.625 mm collimation. The voltage is 120kV for thorax, cervical spine, abdomen-pelvis and cerebrum and 140kV for skull base. Rotation time is 0.8 seconds for neck, abdomen-pelvis and thorax. Rotation time is 1 second on average for CT brain and 0.5 seconds when associated with patient head movement. X-ray tube current varied according to patient size and weight. The current range and corresponding scan type are: 80-335 mA cerebrum; 380-380 mA skull base; 120-220 mA abdomen-pelvis; 100-280 mA cervical spine and 60-160 mA thorax. Minimum and maximum current levels at CT brain are determined using automatic tube current modulation based on one anteroposterior and one lateral pilot scan. Noise index is 3.1 for brain, 35.36 for abdomen-pelvis and 41.42 for thorax respectively.

### 3.4 Radiation dose data extraction

Radiation dose reports are generated using dose measurements calculated from a CT scanner dose area product (DAP) meter. The output is in dose-length product (DLP). The DAP meter is calibrated every six months by a GE engineer. Radiation dose information was extracted from the radiation dose reports for each patient. A typical dose report is pictured in Figure 1 below. Dose reports from the GE LightSpeed VCT-XTE are automatically transmitted via the PACS system along with the associated patient imaging. This allowed rapid access to the relevant dose information. For each scan category the scan type, CTDIvol, scan range, DLP and patient age were recorded in the database. The work was initially carried out by a final year medical student and data cleaning was subsequently conducted by the senior UCC lecturer in radiology before ED calculation was undertaken.

**Figure 1.**
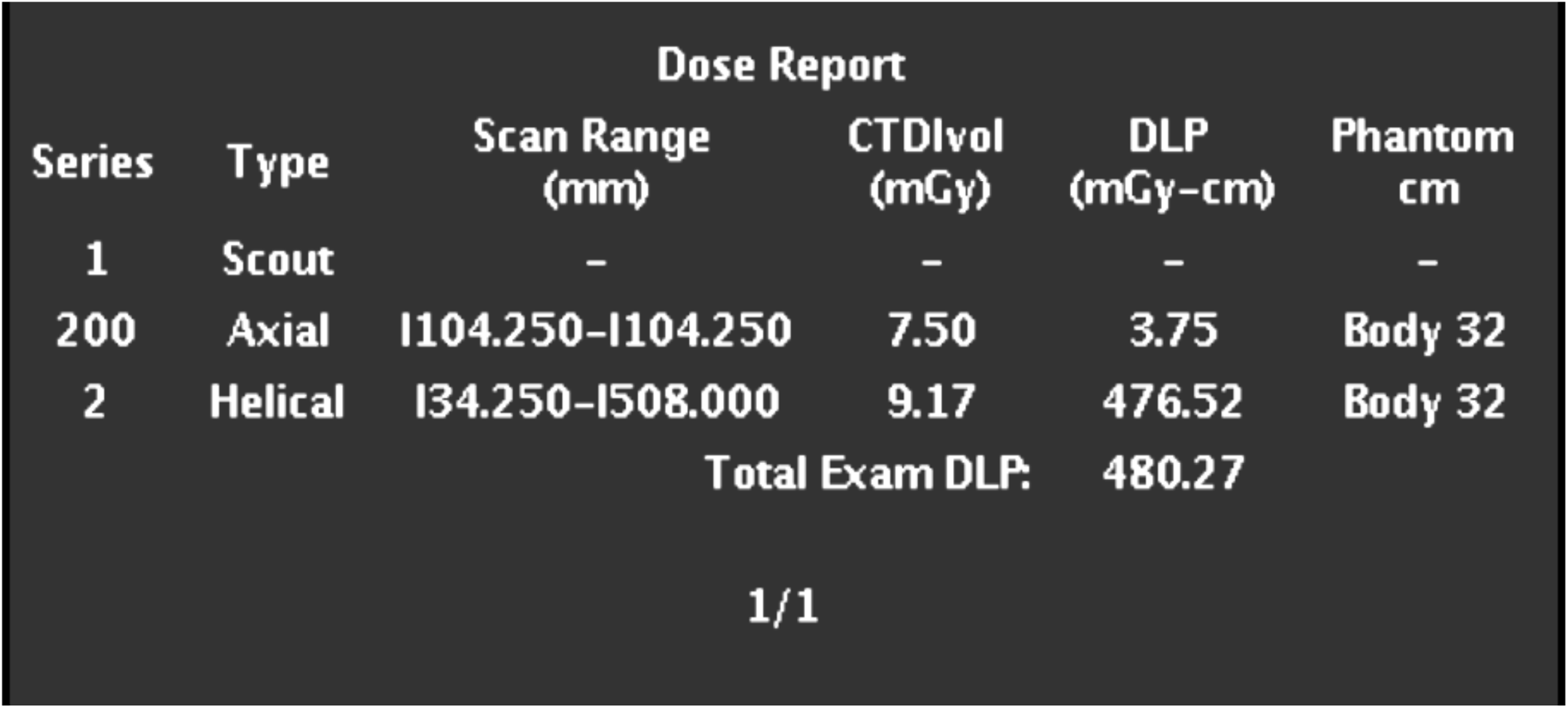
Sample dose report from GE LightSpeed VCT-XTE.

### 3.5 Effective Dose calculation

Before calculating the ED, it was necessary to choose a robust method of ED estimation. In studies comparing effective dose calculated using Monte Carlo methods and DLP-based conversion coefficients, it was found that the two methods do not differ significantly (15, 21, 26). In a study of seven CT protocols, DLP conversion methods based on IRCP 60 weighting factors tended to underestimate ED in comparison to organ dose methods by a mean percentage variation of less than 6% (15). These differences in estimates of ED suggested the need to reassess DLP conversion coefficients by adopting the new ICRP 103 weighting factors particularly for scans over the breast and thyroid. DLP-based methods are also more reflective of the practical working conditions and time constraints of a radiology department compared to the more computationally complex Monte Carlo methods (27). Calculation of ED from DLP allows comparison of ionising radiation exposure with other radiological imaging modalities and the natural background radiation exposure in Ireland of 4 mSv per annum (6, 28).

DLP conversion factors based on the new IRCP 103 estimates from the work of Huda at al. were used to calculate the ED for the current study (29). Conversion factors were based on examinations performed at 120 kV in 32cm acrylic phantoms for all body examinations (including neck) and in 16cm acrylic phantoms for all head examinations. Results for conversion factors had been rounded to two significant figures. Factors for chest, abdomen-pelvis and neck DLPs were 0.02, 0.015 and 0.011 uSv/mGy.cm respectively. The head DLP conversion factor was 0.0024 uSv/mGy.cm. Factors were applied according to image type for all DLPs and ED results were exported to a new Microsoft Excel 2007 spreadsheet for further statistical and mathematical manipulation.

### 3.6 Acquisition of local and national DRLs

Published DRLs specific to Ireland were not available at the time of writing. However, the UCD School of Medicine is currently surveying 34 CT scanners in Ireland as part of a national project and has calculated DRL values (75th percentile) at both a local and national level. DRLs specific to the GE LightSpeed CT system at CUH (Table 1) and at a national level (Table 2) were kindly made available to the author.

**Table 1.** DRLs for GE LightSpeed CT, CUH

| CT Procedure | DLP (mGy cm) | Effective Dose (mSv)<sup>1</sup> |
| --- | --- | --- |
| Brain | 980 | 2.06 |
| Cervical Spine | 520 | 3.07 |
| Routine Chest | 600 | 8.4 |
| Routine Abdomen – Pelvis | 650 | 9.75 |

<sup>1</sup>Using DLP to convert to effective dose, conversion coefficients from NRPB W67, *Doses from Computed Tomography (CT) in the UK - 2003 Review*.

**Table 2.**
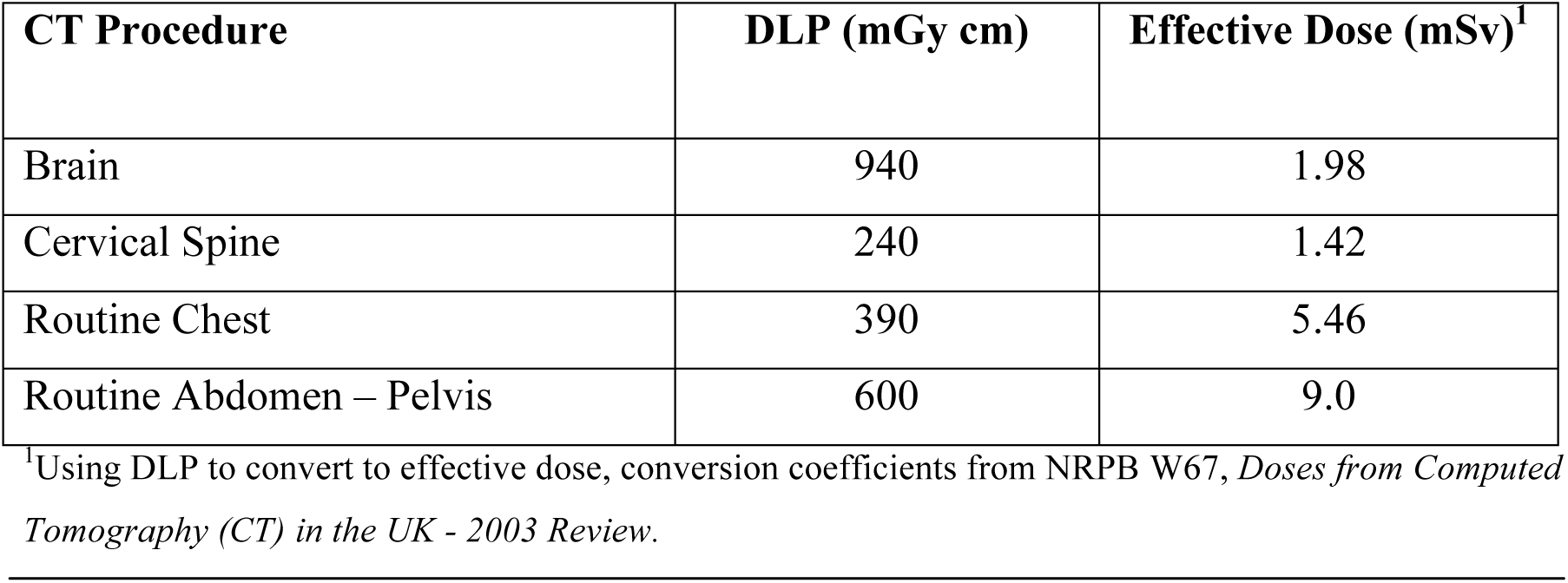
Irish National DRLs

| CT Procedure | DLP (mGy cm) | Effective Dose (mSv)<sup>1</sup> |
| --- | --- | --- |
| Brain | 940 | 1.98 |
| Cervical Spine | 240 | 1.42 |
| Routine Chest | 390 | 5.46 |
| Routine Abdomen – Pelvis | 600 | 9.0 |

<sup>1</sup>Using DLP to convert to effective dose, conversion coefficients from NRPB W67, *Doses from Computed Tomography (CT) in the UK - 2003 Review*.

### 3.7 Comparison with international effective dose values

Literature was reviewed through Pubmed and EBSCO via the UCC library network using the terms ‘effective dose’, ‘CT’, ‘computed tomography’, ‘radiation dose’, ‘patient dose’ and ‘dose length product’. Additional papers were accessed by cross referencing bibliographies from retrieved articles. The search was limited to results from adults in the time period from 1989 to September 2011. Dosimetric quantities were extracted for thorax, abdomen-pelvis, cervical spine and head examinations. ED and its variation among studies was assessed with respect to country or area of origin, year of publication and CT system studied (single slice versus multislice). Reported dosimetric quantities differed between studies with ED being the most commonly quoted measure. DLP was converted to ED, where appropriate, using IRCP publication 103 weighted conversion factors (n=3). The results are presented in Table 3.

**Table 3.**
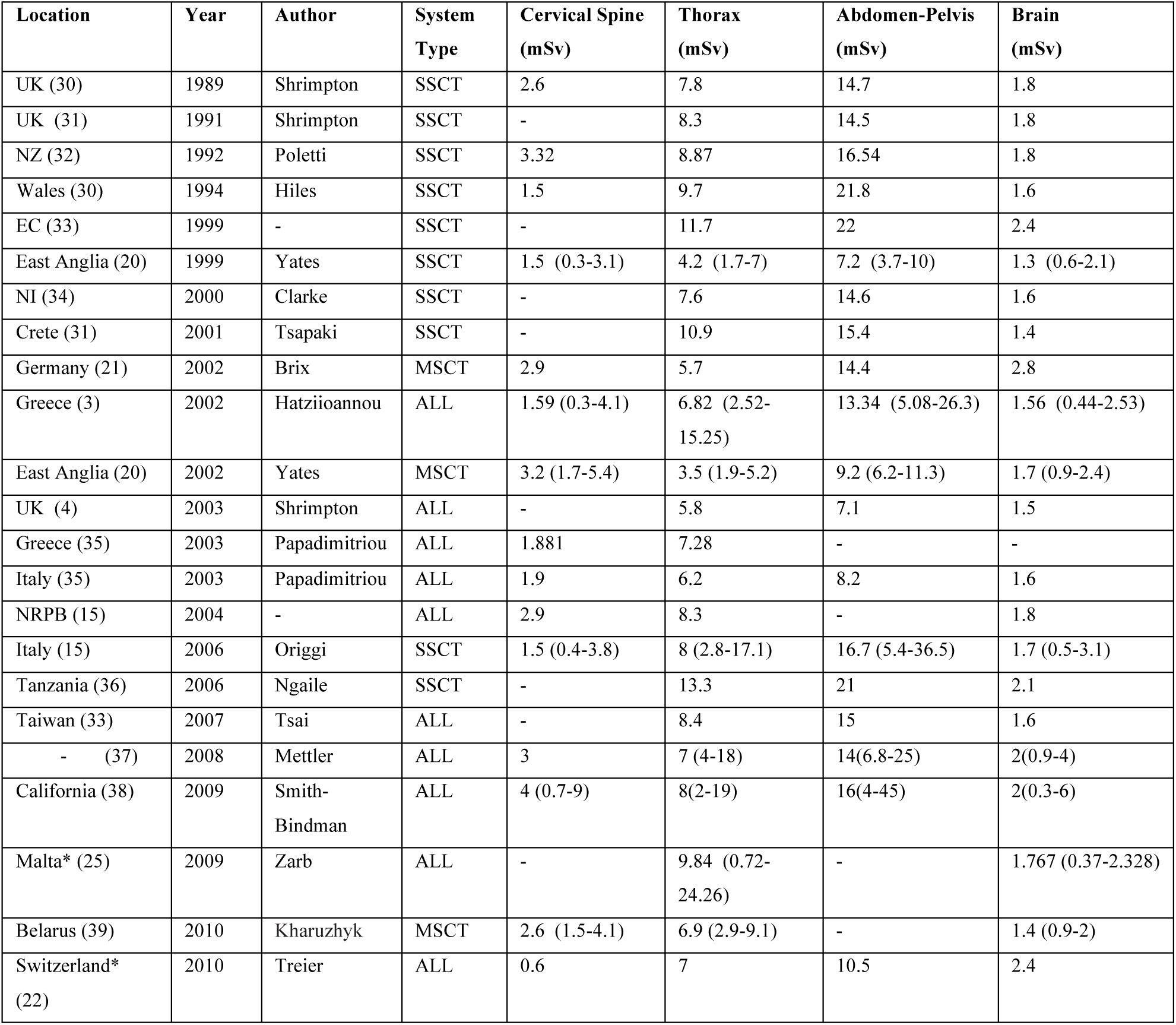
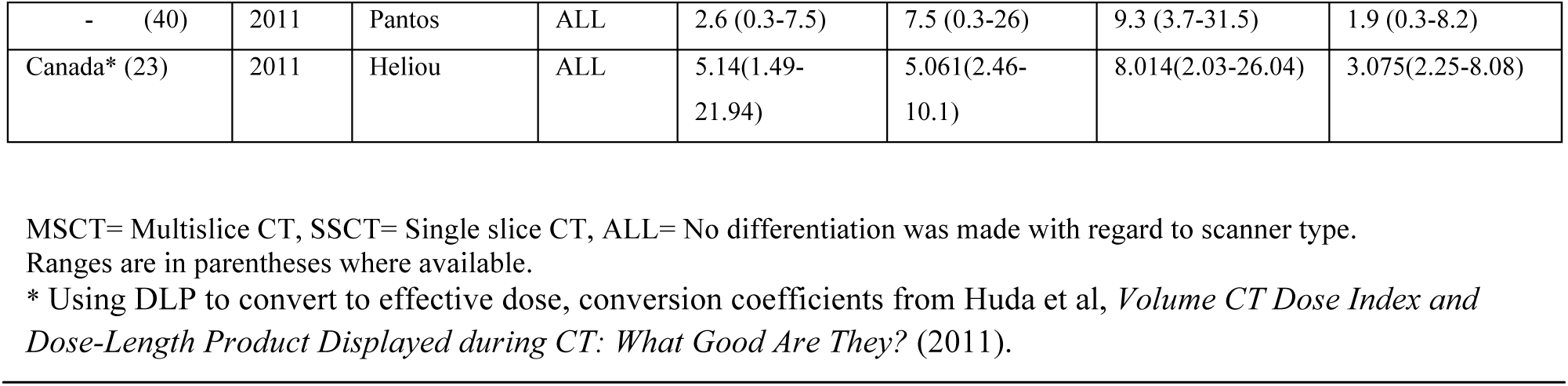
Summary of published international effective dose means (1989-2011)

**Table 4.**
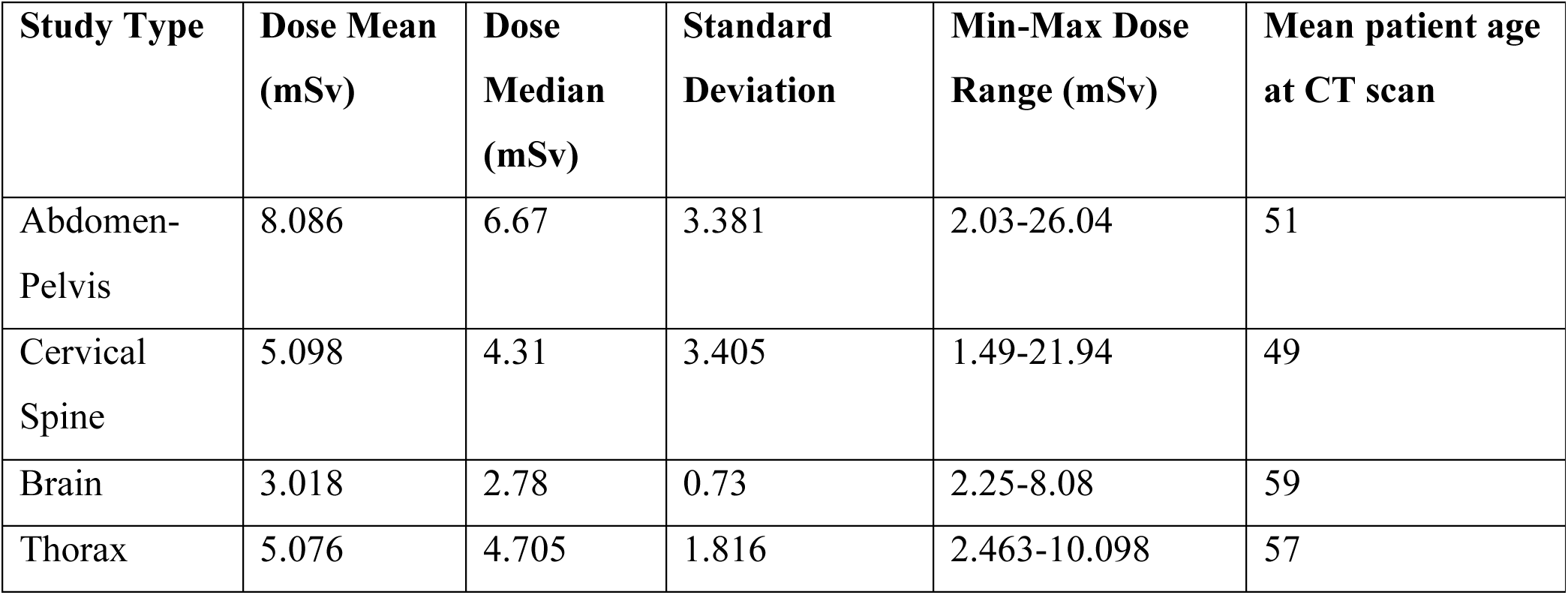
Summary of ED values and mean patient age for four scan types

| Study Type | Dose Mean (mSv) | Dose Median (mSv) | Standard Deviation | Min-Max Dose Range (mSv) | Mean patient age at CT scan |
| --- | --- | --- | --- | --- | --- |
| Abdomen-Pelvis | 8.086 | 6.67 | 3.381 | 2.03-26.04 | 51 |
| Cervical Spine | 5.098 | 4.31 | 3.405 | 1.49-21.94 | 49 |
| Brain | 3.018 | 2.78 | 0.73 | 2.25-8.08 | 59 |
| Thorax | 5.076 | 4.705 | 1.816 | 2.463-10.098 | 57 |

### 3.8 Statistical analysis

Statistical analysis took place after consultation with a qualified statistician and epidemiologist at the UCC School of Medicine MX5090 statistical consultation session. It was advised that statistical results be displayed and examined by graphical and tabular representation to allow for visual comparison of results from CUH with previously published results.

Data was exported from Microsoft Office Excel 2007 to SPSS version 18.0 for Windows (IBM Corporation, NY, US). Histograms were generated for the demonstration of local distribution of radiation dose at CT scan on SPSS. The calculation functions of Microsoft Excel were used to generate range, mean, median and standard deviation of ED for each study category. Further charts were created on Microsoft Excel based on data collected from review of the literature for reference ranges. This information was plotted using point values and ranges where they were available in the literature. Correlation between CTDIvol, Total Scan Length and ED was calculated using linear regression analysis.

### 3.9 Study ethics

Expedited ethical approval was granted for this project in July 2011 by the Clinical Research Ethics Committee of the Cork Teaching Hospitals (Appendix A).

### 3.10 Study timeline

Data collection and calculation of effective dose was carried out in the radiology department of CUH between July and November 2011.

## Section 4: Results

### 4.2 Summary of ED for each of four scanner categories at CT in CUH

### 4.2 Comparison with National and Local DRLs

The ED distribution for each category type was plotted on a histogram, and all studies were compared to both local and national DRLs. Effective dose frequency at CT abdomen-pelvis was 28.5% above the national DRL (9 mSv) and 26% above the local DRL (9.75 mSv). Effective dose at CT thorax was 3.5% greater than local DRL (8.4mSv) and 42.5% greater than the national DRL (5.46 mSv). Effective dose at CT brain was 100% above both the local (1.98 mSv) and national (2.06 mSv) DRLs. CT cervical spine effective dose was 79% greater than the local DRL (3.07 mSv) and 100% greater than the national DRL (1.42 mSv).

### 4.3 Comparison with international mean doses

The mean ED values for CT cervical spine, thorax, abdomen-pelvis and brain scans were analysed by comparison with mean values cited in international studies of CT dose as outlined in Table 3. A notable individual study is Mettler et al’s *Effective Doses in Radiology and Diagnostic Nuclear Medicine: A Catalog* (2008). This seminal work is the most cited paper in the literature on radiation dose and comparisons were made on an individual basis with this key study.

The mean ED at CT abdomen-pelvis scan of 8.041 mSv is 37.8% less than the average of all other studies (including Mettler et al) (n=20) at 13mSv and 42.24% less than the average dose quoted by Mettler et al at 14mSv. The mean ED of CT thorax of 5.061 mSv 33.73% less than the average of all other studies (n=25) at 7.65mSv and is 27.57% less than the average dose quoted by Mettler et al at 7mSv. The mean ED of CT brain of 3.075 mSv is 52.36% greater than the average of all other studies (n=24) at 1.84mSv and is 54% greater than Mettler et al’s doses at 2 mSv. The mean ED of CT cervical spine of 5.14mSv is 85.1% greater than the average of all studies (n=17) at 2.75mSv and 69.8% greater than Mettler et al’s doses at 3mSv. The results are displayed in Figures 6-9.

**Figure 2.**
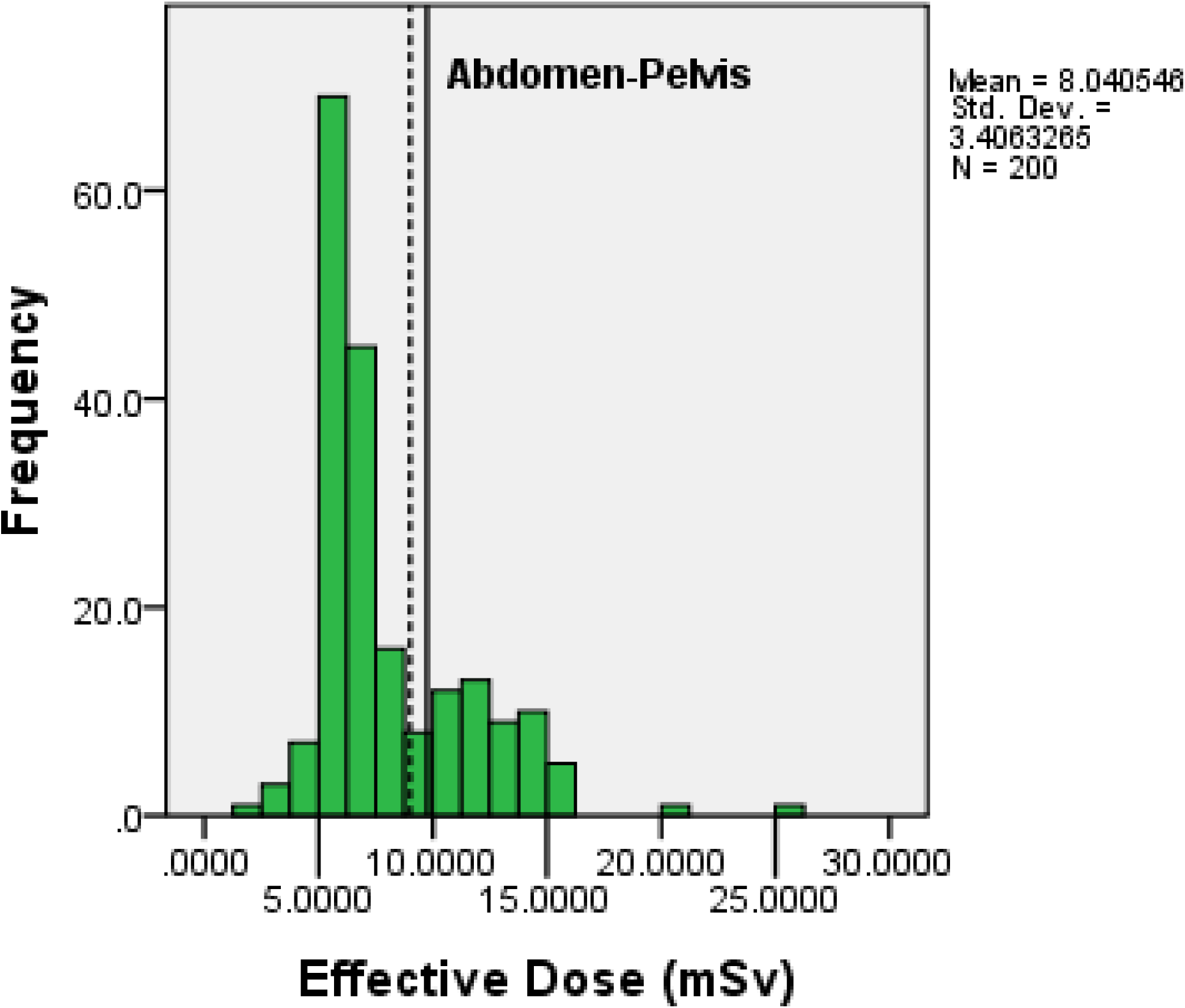
Frequency of effective dose at CT abdomen-pelvis routine. National DRL (broken reference line) is at 9 mSv. Local DRL (solid reference line) is at 9.75 mSv

**Figure 3.**
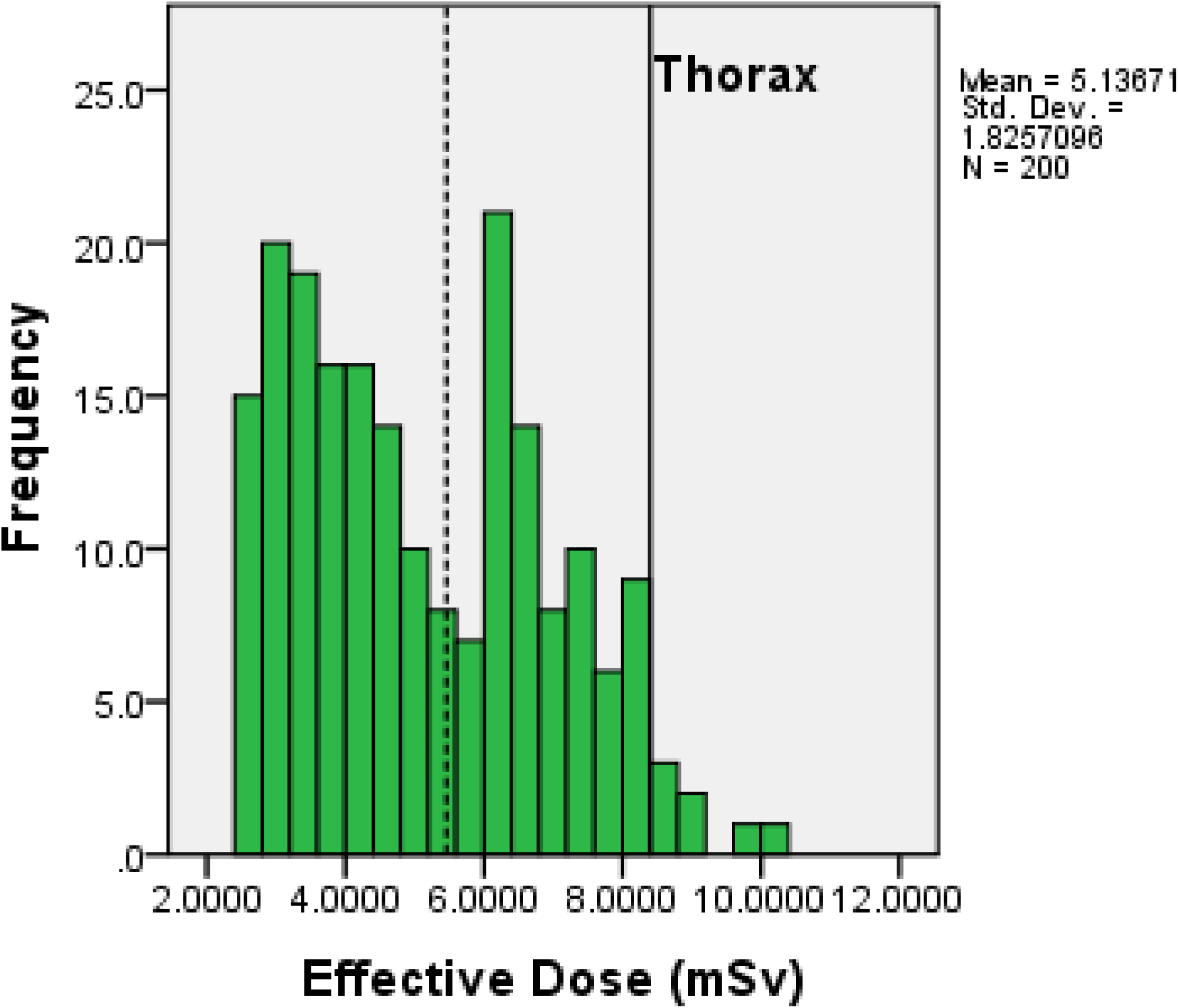
Frequency of effective dose at CT Thorax with contrast routine. National DRL (broken reference line) = 5.46 mSv. Local DRL (solid reference line) = 8.4mSv.

**Figure 4.**
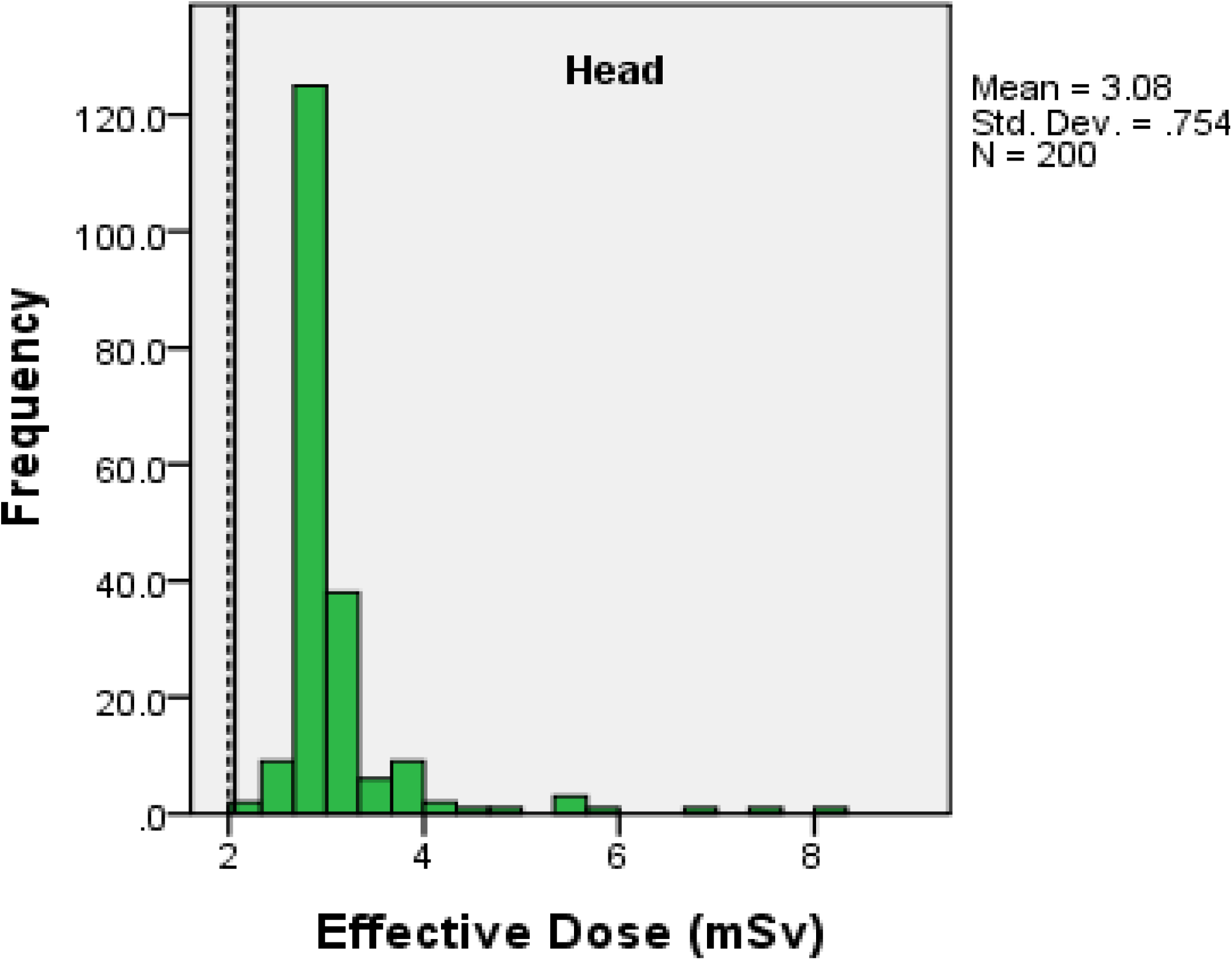
Frequency of effective dose at non-contrast CT Brain routine. National DRL (broken reference line) = 1.98 mSv. Local DRL (solid reference line) = 2.06 mSv.

**Figure 5.**
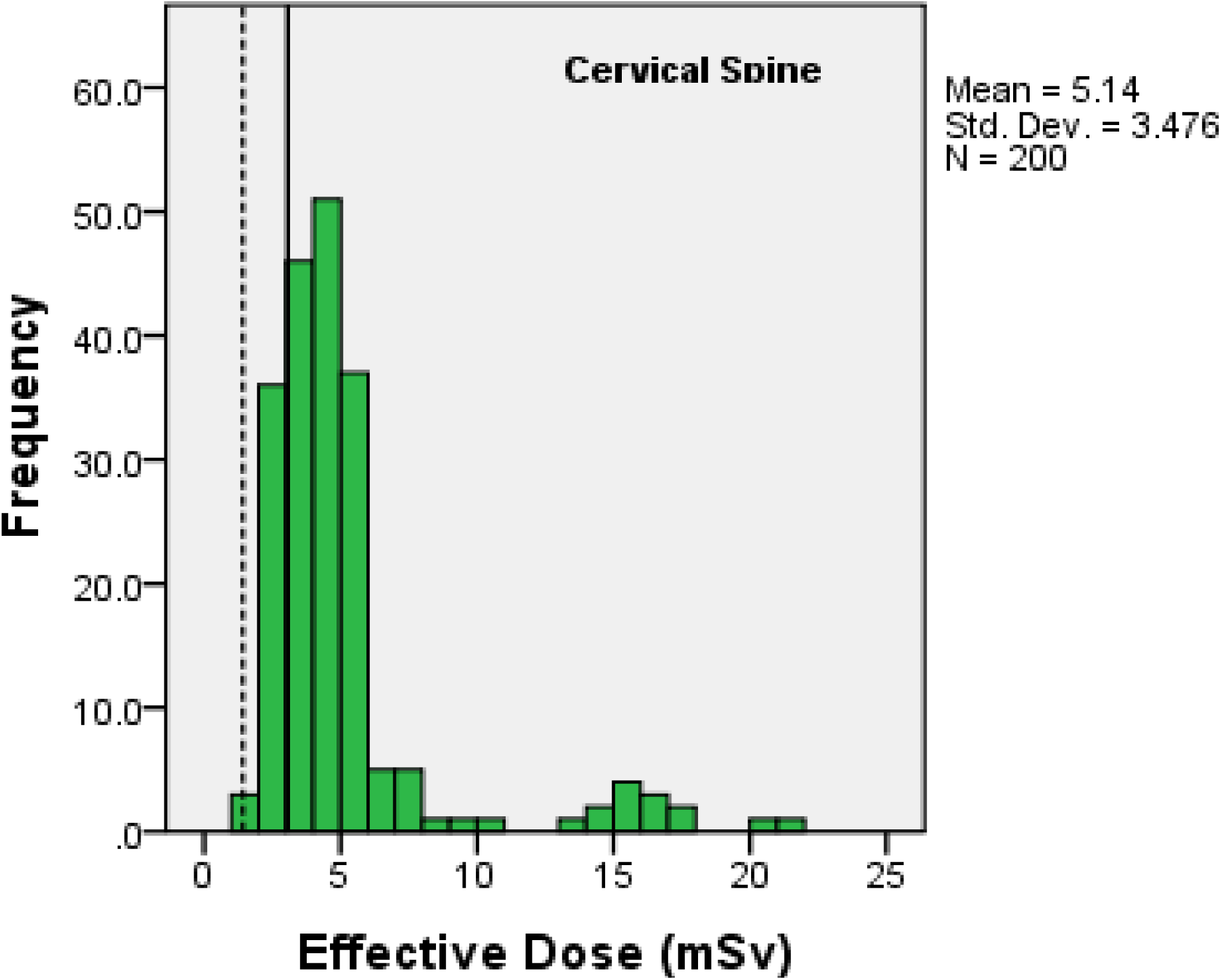
Frequency of effective dose at CT Cervical Spine routine. National DRL (broken reference line) is at 1.42 mSv. Local DRL (solid reference line) is at 3.07 mSv

**Figure 6.**
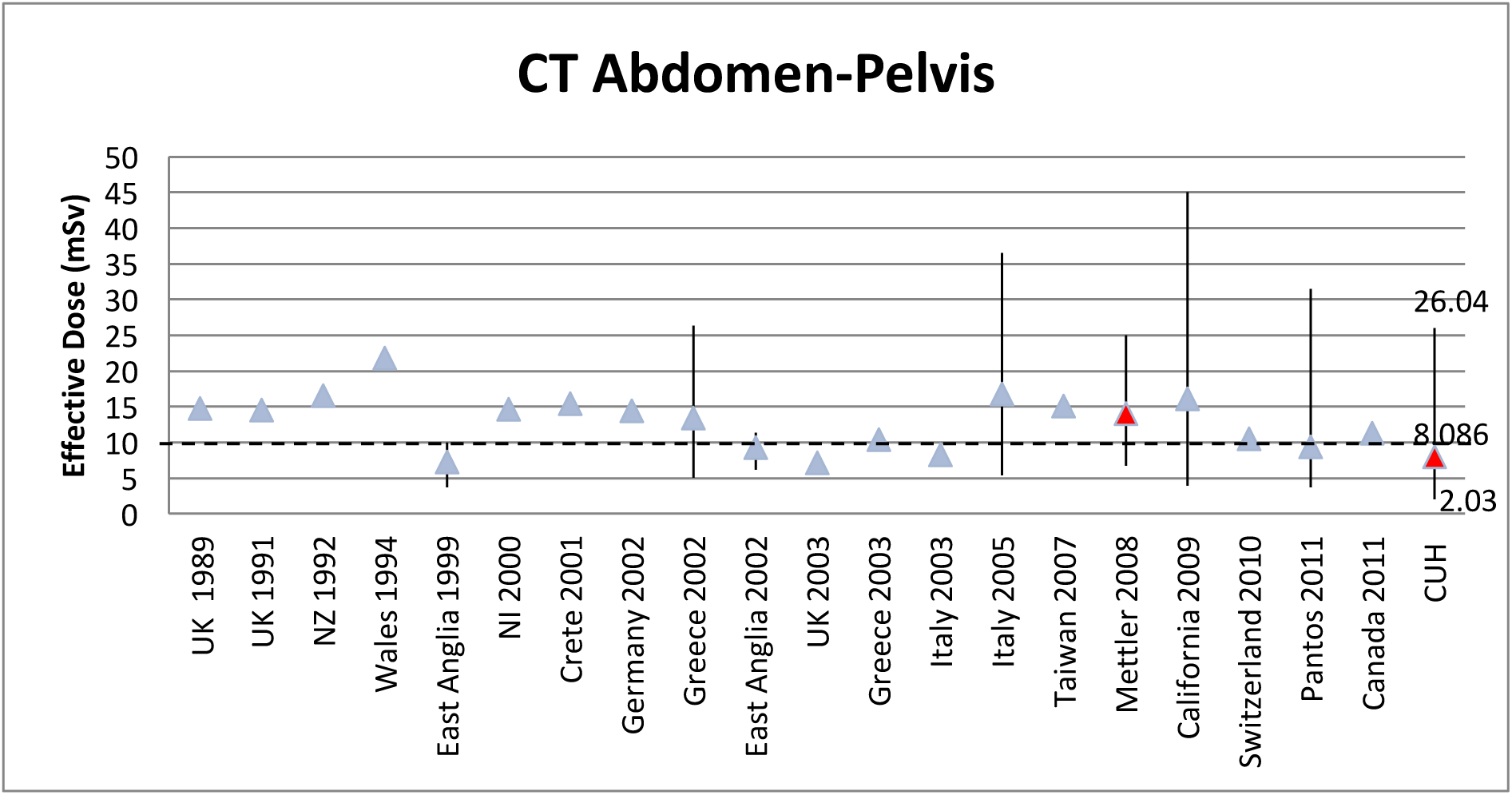
Comparison of mean point value and range values of CT Abdomen-Pelvis at CUH with International levels. The average value of all mean point values of 13 mSv is indicated by a broken line.

**Figure 7.**
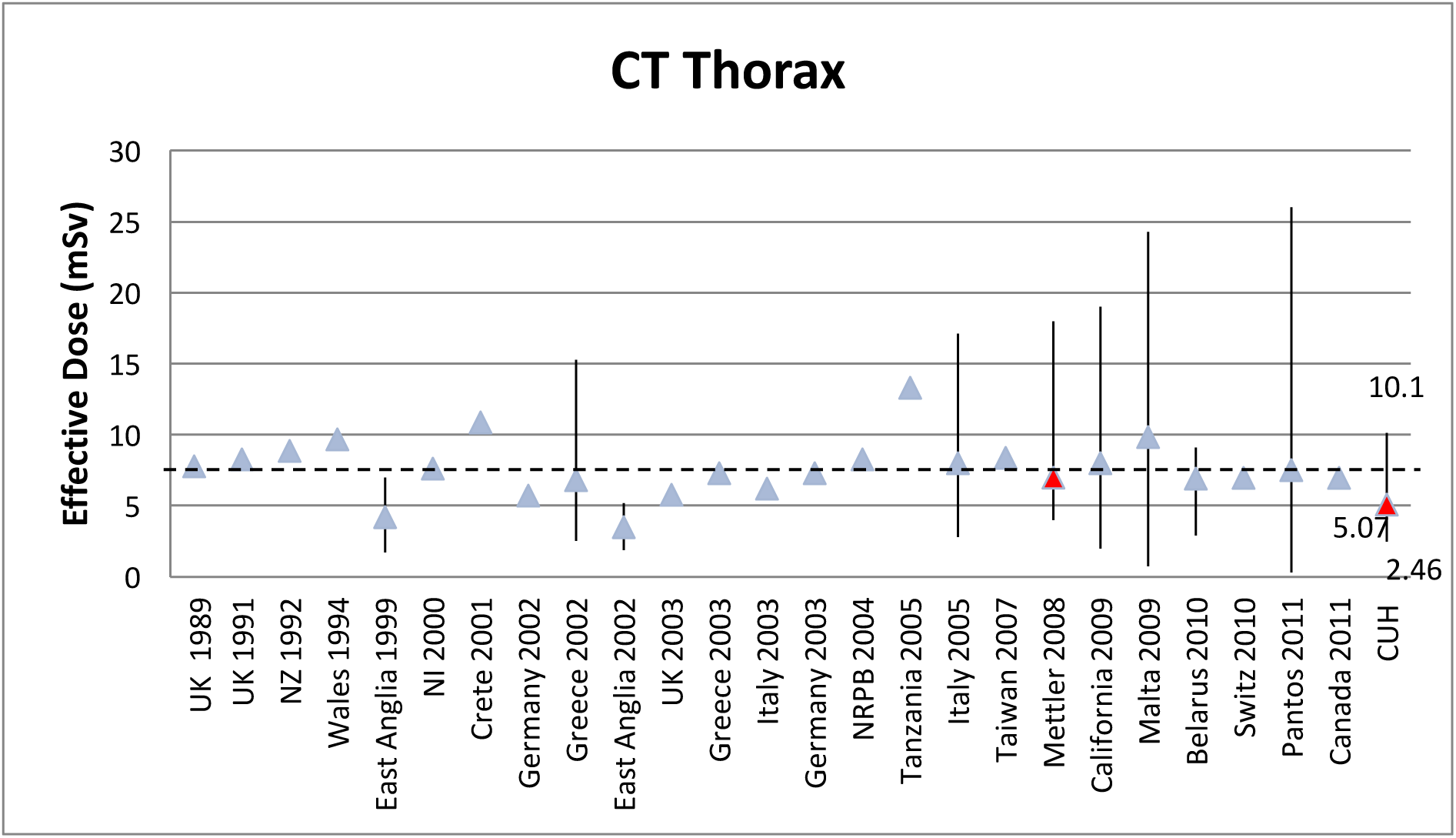
Comparison of mean point value and range values of CT Thorax at CUH with International levels. The average value of all mean point values of 7.65 mSv is indicated by a broken line.

**Figure 8.**
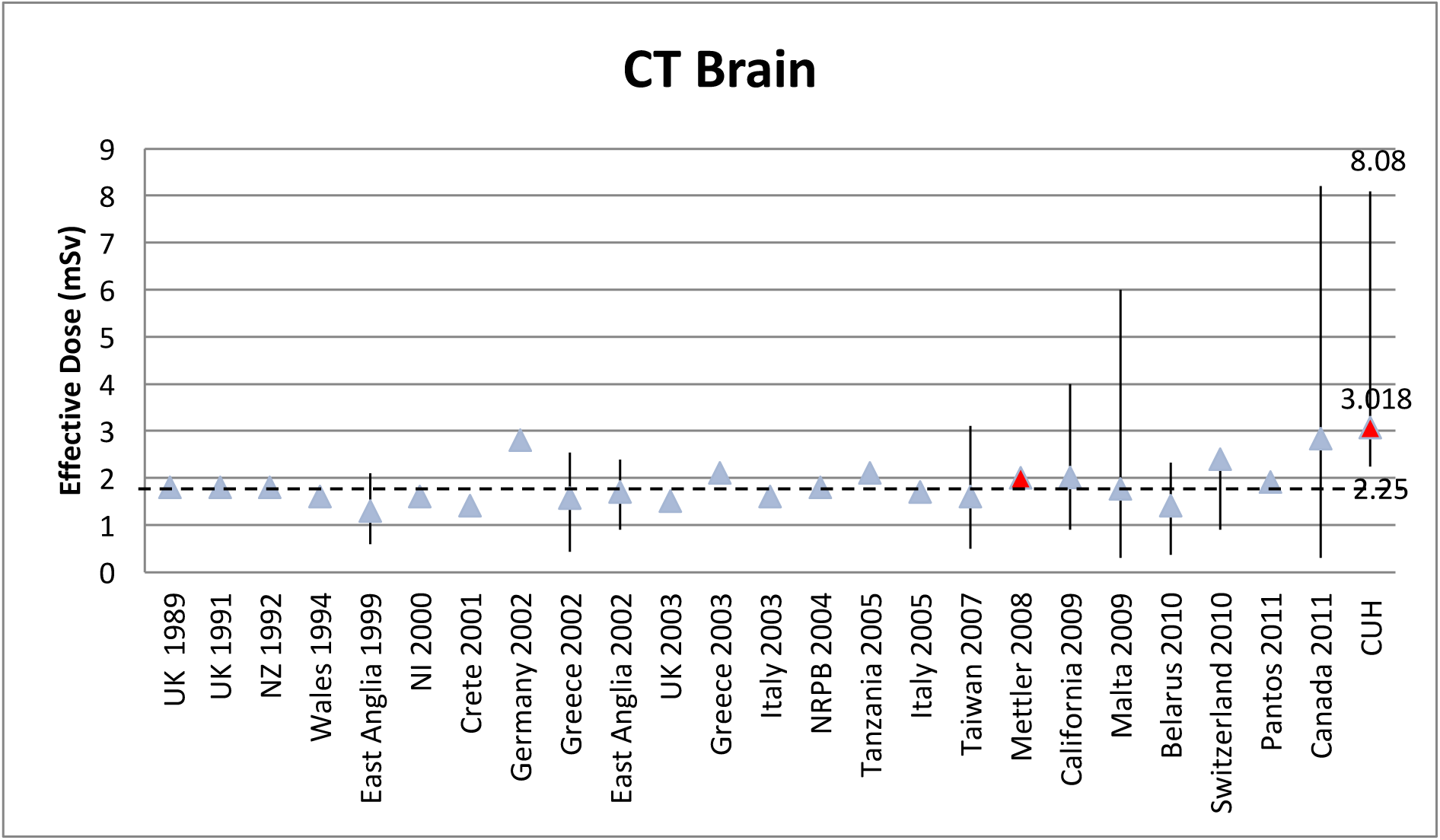
Comparison of mean point value and range values of CT Brain at CUH with International levels. The average value of all mean point values of 1.84 mSv is indicated by a broken line.

**Figure 9.**
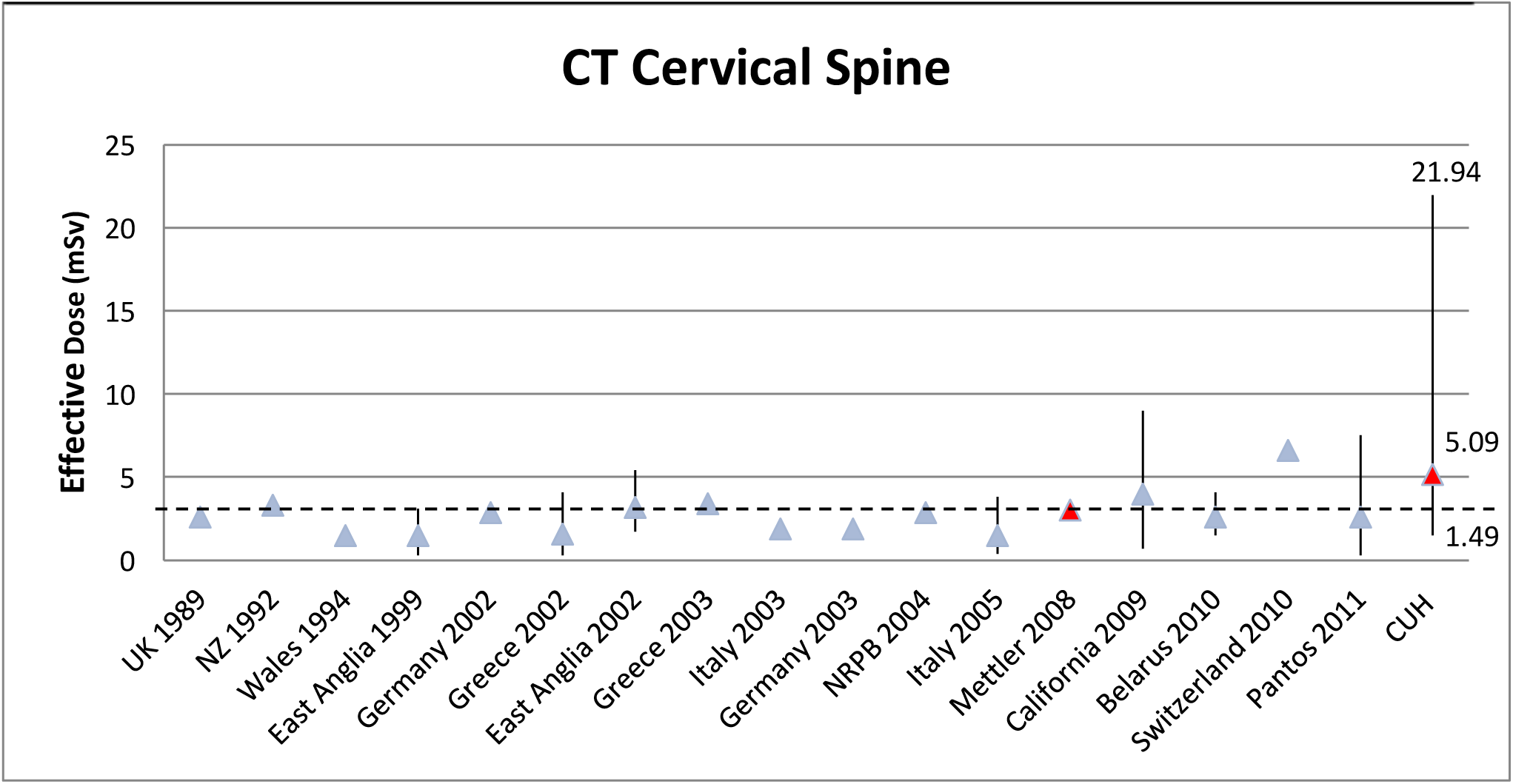
Comparison of mean point value and range values of CT Brain at CUH with International levels. The average value of all mean point values of 2.75 mSv is indicated by a broken line.

### 4.4 Contribution of scan length and CTDIvol to ED

The relationship between the scan parameters is described by analysing the correlation between total scan length (TSL), CTDIvol and ED using linear regression analysis. This can be used to elucidate the underlying relationships of the wide variation in ED values by analysis of CTDI and TSL variance.

## Section 5: Discussion

The notable advances in CT technology and applications over the past decade have increased the clinical utilisation of CT but have also generated concerns about individual and population doses of ionising radiation (14). While the benefits of CT exceed the harmful effects of radiation exposure in patients, increasing population doses have raised a compelling case for reduction of radiation exposure from CT (1). This has prompted the implementation of several radiation reduction techniques which focus on the design of dose efficient software and the optimisation of scan protocols (41). To date, these techniques have primarily focused on software optimisation technologies which either improve CT scanner efficiency or increase image quality at low radiation dose (14, 42).

Improving CT scanner efficiency involves modulating tube output in response to patient attenuating factors. This is achieved using x-ray filters and automated tube current modulation. Bow-tie filters may be used to reduce the patient absorbed dose by 50% compared to flat filters (42). Traditionally, fixed tube kilovoltage and ampere protocols have been used in CT imaging which lead to inefficient radiation exposure due to the relative dissociation between radiation exposure and image quality at higher radiation doses (2).

Automated tube current modulation automatically alters the tube current at CT in response to the size and attenuating properties of the body part being imaged in the x-y plane (angular modulation) or along the z-axis (z-axis modulation) (43). Compared with fixed tube current ATCM can lead to a reduction of 32% in tube current product in up to 87% of examinations (44). Radiation dose reductions with ATCM of 20-40% have been noted when image quality is appropriately specified (45).

Methods of maintaining image quality at low radiation doses have been developed with varying success. These methods include post-processing CT images with Noise Reduction Filtration (NRF) software and improving on the computational power of image reconstruction algorithms by using iterative reconstruction in place of filtered back projection (FBP). It has been shown that NRF is effective in reducing image noise by 30-60% predominantly along the line of greatest attenuation but it has also been documented that there is compromised diagnostic acceptability and lesion conspicuity, particularly in abdominal and chest imaging (42).This method has been superseded by iterative reconstruction which represents the most exciting radiation reduction technology available today. This technology has the potential to improve image quality in scans with reduced tube currents by the reconstruction of images with lower image noise compared to the simpler mathematical model used in FBP techniques (45, 46).The most common iterative reconstruction algorithm in use is Adaptive Statistical Iterative Reconstruction (ASIR). ASIR selectively identifies and then subtracts noise from an image using information from FBP as the initial building block of the process (46).

The implementation of DRLs is a key element in any radiation reduction strategy. They represent an important protocol optimisation tool which is now endorsed by many professional and regulatory organisations, including the ICRP, United Kingdom Health Protection Agency, American College of Radiology (ACR), European Commission (EC) and the International Atomic Energy Agency (IAEA) (24). When used in non-CT based radiological studies DRLs have lead to significant decreases in radiation doses. UK national dose surveys demonstrated a 30% decrease in typical radiographic doses from 1984 to 1995 and an average reduction of approximately 50% between 1985 and 2000 (24, 47). These reductions reflect improvements in reduction strategies and hardware efficiency over time. However the appropriate use of any dose reduction strategy is directed by DRLs. As well as triggering investigations into a particular study type when a reference dose is exceeded, DRLs play a role in providing a benchmark for the overall implementation and progress of modern dose reduction technologies in a centre.

The results of the current study were attained in a centre where low dose imaging protocols have been used since the installation of the system studied in 2009. The dose reduction technologies in place for all study categories audited include bow-shaped filters, ATCM and ASIR. The current study is unique in that it is the first to apply national DRLs specific to Ireland. This has allowed a first insight in the progress made by the application of dose reduction technologies in this country. The results of the study will serve as useful markers for future performance in comparison to both national and local values after further dose reduction measures are applied. The wide disparity between local and national DRLs, seen particularly at CT thorax (5.46 mSv local DRL, 8.4mSv national DRL) is indicative of the wide variety of CT scanners in use in Ireland.

The results document that ED for all four scan categories exceeds the national DRL. Mean ED measurements were crosschecked by comparison with international published values. The ED at CT abdomen-pelvis and CT thorax compared favourably with international published data. Mean ED in both studies was greater than 35% less than the international average dose. However, abdomen-pelvis and thorax studies exceeded both local and national DRLs. ED at CT brain and CT cervical spine was 54% and 85% greater than the international average dose respectively. These results were substantiated by both studies being significantly above local and national DRLs. CT brain was 100% greater than both the local (1.98 mSv) and national (2.06 mSv) DRLs. CT cervical spine ED was 79% greater than the local DRL (3.07 mSv) and 100% greater than the national DRL (1.42 mSv). It is to be expected that all studies would perform poorly in respect to DRLs given the absence of prior reference levels to guide clinical practice in Ireland. However the degree to which CT brain and cervical spine studies exceed these levels is surprising.

One limitation of the current study was the lack of data to elucidate underlying causes of excess dose. Scan-specific protocol data when compared to patient demographic data such as patient body mass index, obtained on a chart review, would outline settings where patients were unnecessarily overexposed. Another constraint of a student project is the absence of CT image interpretation. This level of study does not allow observations and recommendations to be made on CTDIvol levels. Nevertheless, given that all studies included were for diagnostic use, the study population was representative of all patients undergoing CT in the centre being studied. The results presented herein are therefore relevant.

An insight into one possible cause of excess dose was given by TSL correlation as outlined in Table 5. In scans were ED was greater than the international average (CT brain and cervical spine), the variation in ED was greater than 60% due to variation of total scan length. With helical CT scanning there is a tendency to increase the area of coverage which increases the effective dose to the patient (1). Scanning beyond anatomical limits in the context of abdomen and pelvis imaging has been demonstrated to be a common practice with 97% of cases having extra images above the diaphragm and 94% having extra images below the symphysis pubis in a published study (2, 48). Scanning beyond anatomical limits has been found to increase radiation dose by as much as 56% with little additional diagnostic information being obtained in both CT abdomen and CT thorax studies (48, 49).

**Table 5.** Correlations of TSL variance with ED variance across four scan types

| Scan type | Correlation ($r^2$ value) | r value | p value |
| --- | --- | --- | --- |
| Brain | 62% | $r=0.789$ | $p<0.01$ |
| Cervical Spine | 69% | $r=0.833$ | $p<0.01$ |
| Abdomen-Pelvis | 13% | $r=0.363$ | $p<0.01$ |
| Thorax | - | $r=0.083$ | $p=0.243$ |

**Table 6.** Correlations of CTDIvol variance with ED variance across four scan types

| Scan type | Correlation ($r^2$ value) | r value | p value |
| --- | --- | --- | --- |
| Brain | 52% | $r=0.727$ | $p<0.01$ |
| Cervical Spine | 68% | $r=0.829$ | $p<0.01$ |
| Abdomen-Pelvis | 71% | $r=0.844$ | $p<0.01$ |
| Thorax | <1% | $r=0.163$ | $p<0.05$ |

Many factors influence the radiation dose associated with CT imaging. In order to lower radiation dose effectively, dose reduction must be carried out in a variety of ways prioritised by the institution which complement advances in modern radiation lowering technologies (2). This involves a multidisciplinary approach requiring careful planning and execution of CT protocols by technologists, medical physicists and radiologists as well as appropriate referral and consideration of low or radiation free alternatives by medical physicians.

## Section 6: Conclusions & Recommendations

In Ireland, no other study has attempted to analyse and audit ED at CT using DRLs. This first application of newly available local and national standards has revealed a significant optimisation potential for effective dose at CT in the centre studied. This has been noted, in particular, for examinations of the head and cervical spine.

Further studies need to investigate the underlying causes of the excessive dose noted in studies involving the head and cervical spine. A review of imaging protocols and patient factors will need to be carried out to identify areas where radiation dose reductions need to take place and to guide changes to the protocols where necessary. As with any audit process, a future audit will be required to gauge improvements in performance with respect to DRLs (50).

There is a clear case for more extensive research in this area in Ireland. Moreover, there is a compelling argument for further investigation into other scan types in the centre studied as the full, published version of local and national DRLs becomes available. Local DRLs will also require future adjustment as newer more advanced reduction technologies become available and are put into use.

## Data Availability

Dataset for the study is available on request.

## Terminology & Abbreviations

ATCM: Automated tube current modulation
ASIR: Adaptive statistical iterative reconstruction
CT: Computed tomography
CTDIvol: Computed tomography dose index (volume)
CUH: Cork University Hospital
DAP: Dose-area product
DLP: Dose-length product
DRL: Diagnostic Reference Level
ED: Effective dose
FBP: Filtered back projection
GE: General Electrical
HSE: Health Service Executive
IRCP: International Commission on Radiological Protection
MDCT: Multidetector Computed Tomography
mGy: milliGrays
MSCT: Multislice Computed Tomography
mSv: milliSieverts
NRF: Noise Reduction Filtration
PACS: Picture archiving and communication system
SSCT: Single slice computed tomography
TSL: Total scan length
US: United States

## Appendix A

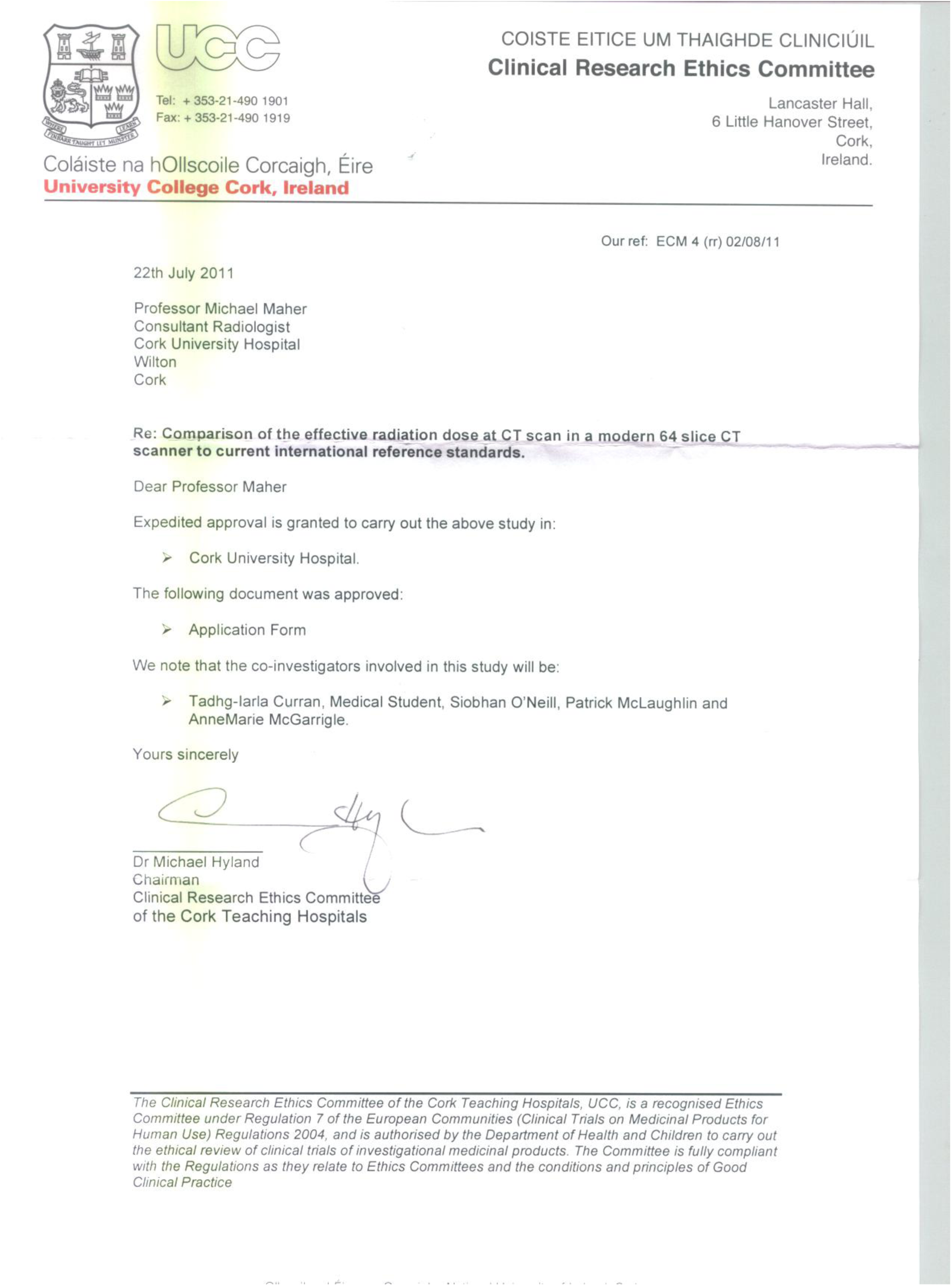

